# Emergency Department Length of Stay, Employment Status, and Subsequent Risk of Hospitalisation: A survival analysis of linked administrative records in England

**DOI:** 10.64898/2026.10.05.26364765

**Authors:** Hannah Aston, Vahé Nafilyan, Adrian Boyle, Daniel Ayoubkhani

**Affiliations:** National Statistician’s Analysis Unit, Office for National Statistics, Newport, UK; London School of Economics and Political Science; Cambridge University Hospitals Foundation Trust

## Abstract

**Objectives:** To investigate the association between time spent in NHS Type 1 emergency departments (EDs, consultant led 24-hour services with full resuscitation facilities) and risk of leaving employment and hospitalisation after leaving the department alive up to two years after attendance.

**Methods:** We conducted a retrospective cohort study using linked administrative data from the NHS Emergency Care Dataset, Hospital Episode Statistics, Census 2021, and mortality records. ED length of stay was categorised from under 2 hours to 12 hours or more. Cox proportional hazards models were fitted to estimate the association between ED length of stay and subsequently leaving employment or being hospitalised over a two-year follow-up period, adjusting for demographic, socioeconomic, and clinical characteristics.

**Results:** Our study population comprised 3,347,008 individuals aged 25–64 who attended a Type 1 ED in England between April 2021 and March 2023. For leaving employment, adjusted hazard ratios (aHRs) increased from 1.04 (95% CI 1.03–1.06) for stays of 2 to <4 hours to 1.21 (1.17–1.24) for 12 hours or more, compared with stays under 2 hours. A similar but more pronounced gradient was observed for post-discharge hospitalisation, with aHRs rising from 1.06 (1.04–1.07) at 2 to <4 hours to 1.95 (1.91–1.99) for stays of 12 hours or more.

**Conclusions:** Prolonged time spent in the ED is associated with poorer outcomes among working-age employees, including reduced labour market participation and increased hospital use. These findings suggest that long ED stays may have wider and longer-term implications beyond immediate clinical outcomes, including potentially for national labour markets and economic growth.

## Introduction

The efficiency of Emergency Departments (EDs) is a key measure of overall NHS stability in England (1). Time spent in the ED has increased substantially in recent years, with nearly 13% of attendances exceeding 12 hours spent in the ED (2), and the four-hour standard introduced in 2004 has not been met since 2013, over a decade ago (3). These extended waits and ED crowding can partly be explained by changes in clinical practice and pathways, high inpatient bed occupancy, reduced capacity (4,5), high bed occupancy rates (6,7) and staffing shortages (8). Additional pressures such as the COVID-19 pandemic have further increased the time patients spend in emergency care, resulting in most EDs consistently failing to meet the four-hour standard (9).

Previous studies have demonstrated that these extended lengths of stay (LOS) may have a determinantal impact upon patients both during the attendance (10) and after, including increased mortality rates after leaving ED care (11–15). In one study, delays to inpatient admission in excess of five hours were found to be associated with an increase in all-cause mortality within 30 days of leaving the ED (11). In another, even after socio-demographic and clinical factors are considered, the risk of 30-day-post-discharge death increased with total time spent in A&E after approximately two hours. Compared with patients requiring non-immediate care who spent two hours in A&E, the odds of post-discharge death were 2.1 times higher for those who spent 12 hours in A&E (15).

Increased ED overcrowding and LOS are occurring alongside a rise in the proportion of people in the UK who are economically inactive due to health reasons (increasing by 40% between 2019 and 2025 (16)). There is growing body of evidence on possible links between healthcare waiting times and economic outcomes, though not necessarily in emergency settings. For example, increased waiting times for elective care are significantly associated with labour market outcomes (17,18), with an estimated £2.3 billion increase in aggregate earnings in England if inpatient elective waiting times met national standard levels (18). There is also evidence to suggest that being hospitalised for a major health condition leads to a consistent decrease in earnings and employment rate across a five-year follow-up period (19).

Although prolonged ED stays is linked to worsened clinical outcomes, there is a lack of evidence on the influence of these stays on economic outcomes. Existing research on the relationship between health and labour market participation largely focuses on other parts of the healthcare system, primarily elective care, and has demonstrated a very clear link between deteriorating health andlabour market outcomes, such as employement and earnings. However, none of these studies directly examine whether delays occurring within the ED, one of the most pressured points of the healthcare system (20), play a similar role. As ED LOS continue to rise, this absence of evidence represents a critical gap in understanding the broader social and economic consequences of emergency care delays.

In this study, we investigated the association between time spent in NHS Type 1 EDs (consultant led 24-hour service with full resuscitation facilities) and subsequent post-discharge risk of leaving employment or being hospitalised, using linked administrative data covering most of the population of England.

## Methods

### Study design and setting

We conducted a retrospective, observational cohort study of individuals who attended a Type 1 ED in England between 1^st^ April 2021 and 31^st^ March 2023 (the latest ED data available to us at the time of analysis) and were discharged alive. Follow-up data ran from 1^st^ April 2021 to 31^st^ March 2025 (the latest available employment and hospitalisations data), allowing for a maximum of two years follow-up.

We created a person-level dataset combining NHS England’s Emergency Care Dataset (ECDS) (21) on ED visits with: the 2021 Census; the Hospital Episode Statistics (HES) Admitted Patient Care (APC) and Outpatient (OP) datasets (22); His Majesty’s Revenue and Customs (HMRC) Pay As You Earn (PAYE) records, cleaned and calendarized by the Office for National Statistics (ONS) using established methods (23); and ONS death registration data. The 2021 Census (24) was linked to the NHS Personal Demographics Service (PDS) to retrieve NHS numbers of Census respondents, with a linkage rate of 95.8% (25). The NHS datasets and death registrations were subsequently linked to the Census using NHS numbers. PAYE records were linked to the Census via the ONS Demographic Index (26) with a linkage rate of 96.7% (27). We only included the individuals’ first ED visit during the study period to maintain the independence of observations in the study sample, as done in previous studies (11,15). This first ED visit defines the index date for the study.

### Selection of participants

Our study population included individuals who were aged between 25 and 64 years (i.e. working-age) who: were enumerated at the 2021 Census as usual residents of England; could be linked to the NHS PDS; could be linked to HMRC PAYE RTI via the Office for National Statistics (ONS) Demographic Index; were a payrolled employee on the date of their attendance according to the calendarized PAYE dataset; visited a Type 1 ED at least once between 1^st^ April 2021 and 31^st^ March 2023; had an ECDS record that was populated with a “non-immediate” acuity level (“low”, “standard”, “urgent”, or “very urgent”); and who survived to discharge from the ED or admission to hospital for inpatient care. The analysis only covered patients with non-immediate acuity levels (all acuity levels other than “immediate”), who represent 98.9% of the total number of ED patients, in order to reduce bias from condition severity (28). For our main analysis, we aggregated the non-immediate acuity groups in this way to increase statistical power. In addition, there is likely to be a degree of inconsistency in how acuity levels are coded in different EDs (29). We excluded individuals recorded as deceased on arrival and removed records that we deemed unreliable: departure date occurring before arrival date; missing information on arrival or departure date or location; or recorded time in the ED exceeding 48 hours.

### Exposure

We measured time spent in the ED in terms of patients’ end-to-end LOS (the difference in hours and minutes between the arrival and departure times recorded in ECDS). Discrete groups were then created as follows: “0 to under 2 hours”, “2 to under 4 hours”, “4 to under 6 hours”, “6 to under 8 hours”, “8 to under 10 hours”, “10 to under 12 hours”, and “12 hours or more”.

### Outcomes

The two primary outcomes were leaving employment (that is, discontinuation of being a payrolled employee as indicated by PAYE records) and being admitted to hospital after the ED visit (identified from HES APC records). For each outcome, if patients experienced the event more than once during follow-up, we only considered the time to their first event. For the hospitalisation outcome, only admissions that occurred at least one day after ED discharge were included (i.e. the outcome definition excludes admissions that directly followed the ED visit).

### Follow-up

Each individual’s follow-up time was split into consecutive 90-day intervals, with the final interval truncated at the end of their observed follow-up. Follow-up began on the date of the index ED visit and continued for up to two years. Participants who remained under observation beyond this two-year window were censored at two years to ensure comparability across the study population. For each of the two study outcomes, individuals contributed person-time until the earliest of the outcome of interest or being censored (the earliest of death, reaching the end of the study period on 31^st^ March 2023, or the two-year administrative censoring point).

### Covariates

We adjusted for the following covariates: age and calendar time in each 90-day interval following the index date; chief complaint, Index of Multiple Deprivation (IMD) (30) quintile group of the patient’s address at, care provider and acuity level at the time of the index ED attendance (from ECDS); sex, ethnic group and highest qualification (a snapshot on 21 March 2021 taken from Census 2021); and 22 comorbidities identified from HES APC and OP data over the five years before 21 March 2021 (chosen for consistency with published evidence (15)). For full list of covariates see Supplementary Table 1.

### Statistical analysis

We described the characteristics of the study cohort at baseline and compared covariate distributions between individuals who spent less than four hours versus at least four hours in the ED using standardised differences, with absolute values greater than 10% indicating meaningful imbalance (31).

We produced Aalen-Johansen cumulative incidence curves to explore the unadjusted relationship between time spent in the ED and each of the two outcome measures, accounting for the competing risk of death in each case. We also produced inverse-probability weighted Aalen-Johansen curves to account for potentially different age and sex distributions between waiting time groups.

To estimate the association between time spent in the ED and each of our two study outcomes - leaving employment and being hospitalised after the ED visit - we fitted separate Cox proportional hazards models, each adjusted for the covariates listed above. Because age and calendar time were expected to vary over follow-up, we implemented a 90-day interval-based data structure to incorporate these variables as time-varying covariates. Due to computational limitations, we took a random 20% sample from our study population for our modelling.

For each model, we exponentiated the estimated coefficients to obtain adjusted hazard ratios (aHRs) in order to assess the relative risk of the outcome occurring for each LOS group, when compared to the reference category (0 to under 2 hours). Uncertainty was quantified using 95% confidence intervals (CIs) around the aHRs.

### Secondary analysis

We fitted an adjusted Cox model for the leaving employment outcome as described above but further incorporated a time-varying binary hospitalisation indicator (whether the patient had been hospitalised up to and including each 90-day interval) as a covariate. This analysis was intended to assess the potential role of post-discharge hospitalisation (a surrogate marker of severe ill-health) as a mediator of the relationship between time spent in the ED and the risk of leaving employment.

We also fitted an adjusted Cox model for each outcome with the ‘immediate’ acuity group included in the study population.

### Patient and public involvement

Due to the general nature of our study population (all ED visits for any reason, rather than with specific health conditions) and outcome definition (hospitalisation or loss of employment for any reason), we did not directly involve patients and the public in the study.

## Results

### Characteristics of study subjects

Between 1^st^ April 2021 and 31^st^ March 2023, 3,347,008 individuals attended a Type 1 ED in England while being a payrolled employee and met our study inclusion criteria. See Supplementary Figure 1 for the sample flow diagram. The mean age of the study population at the index ED attendance was 42.2 (SD 10.8) years, 54.7% of individuals were female and 80.6% were White. Mean time spent in the ED was 4.4 hours.

Compared with patients who spent less than the standard LOS of four hours in the ED, patients who spent at least four hours in the ED were more likely to: be older; have attended the ED with a chief complaint of “Airway / breathing”, “Circulation / Chest”, “Gastrointestinal”, “General / minor / admin”, “Genitourinary”, “Neurological” or “Psychosocial / Behaviour change”; and have a recorded acuity level of ‘Urgent’. Only age, the chief complaint groups and acuity groups had an absolute standardised difference above 10%. For all other covariates, the absolute standardised difference between patients who spent less than four hours vs. at least four hours in the ED was less than 10% (Supplementary Table 2).

### Main results

During follow-up, 733,803 individuals in the study population (21.9%) experienced at least one transition out of employment after the ED attendance, and 1,237,273 (37%) were hospitalised at least once following discharge from the ED.

Unadjusted cumulative incidence curves for both outcomes showed a consistent gradient in which longer times spent in the ED were associated with higher risks. By the end of follow-up, the cumulative incidence of leaving employment was 25.5% among patients who spent 12+ hours in the ED versus 20.2% among those who were in the ED for less than two hours (Figure 1). For post-discharge hospitalisation, the cumulative incidence was 55.5% for 12+ hours in the ED versus 31% for less than two hours (Figure 2). See Supplementary Figures 2 and 3 for age- and sex-weighted curves, which show the same gradients as the unadjusted curves.

**Figure 1.**
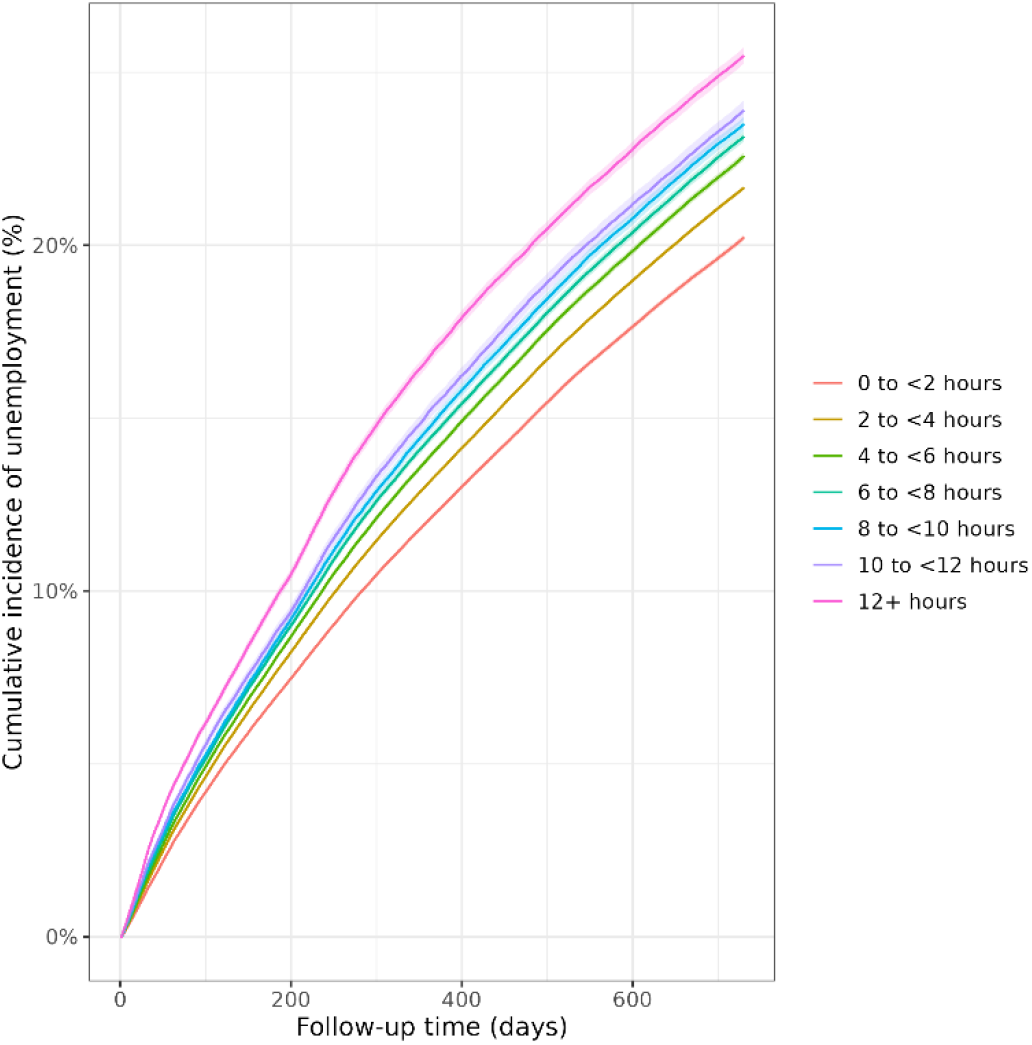
Unadjusted cumulative incidence curves for leaving employment after discharge from the index ED attendance.

**Figure 2.**
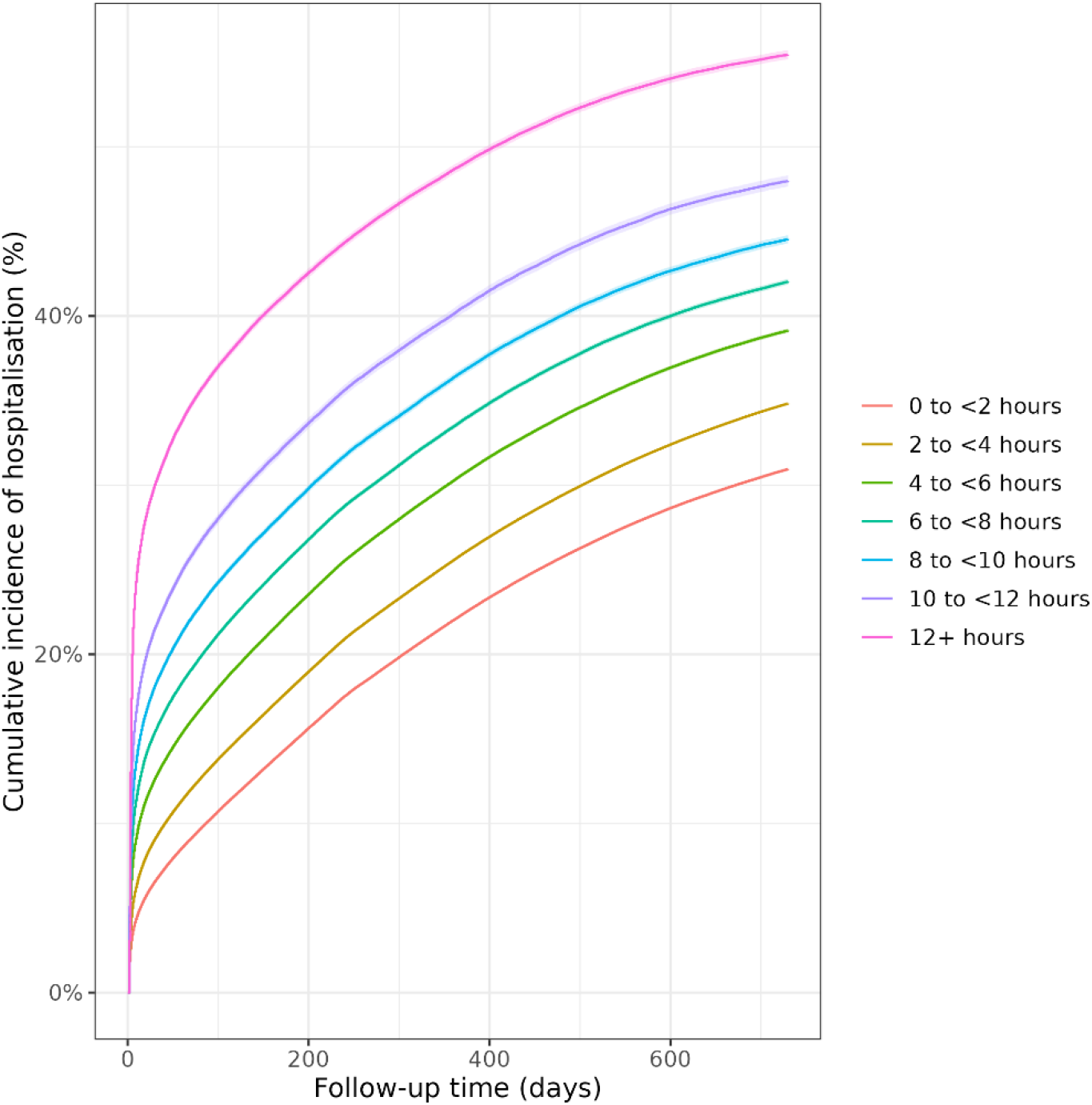
Unadjusted cumulative incidence curves for being hospitalised after discharge from the index ED attendance.

**Figure 3.**
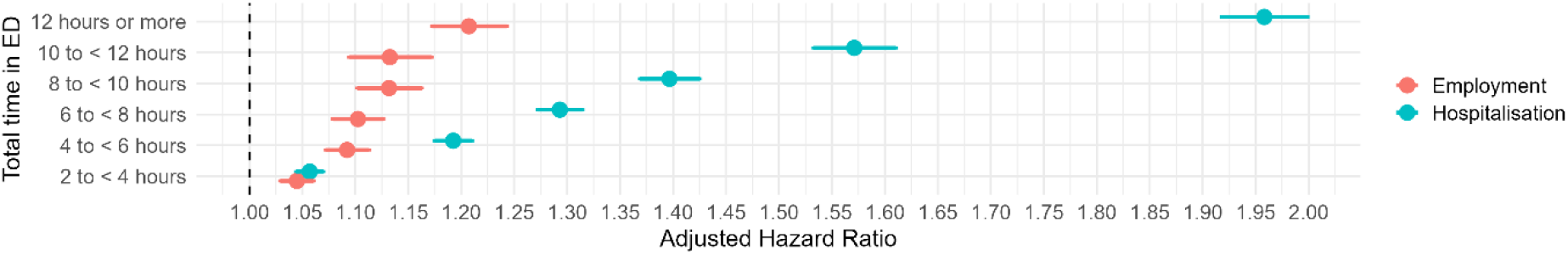
Adjusted hazard ratios for employment and hospitalisation after discharge from the index ED attendance.

### Adjusted models

The aHRs for leaving employment (see Figure 3) increased with total time spent in the ED when compared with the reference group (0 to <2 hours). Individuals waiting 2 to <4 hours had an aHR of 1.04 (95% CI: 1.03 to 1.06), rising to an aHR of 1.21 (95% CI: 1.17 to 1.24) for more than 12 hours spent in the ED.

The aHRs for post-discharge hospitalisation showed a similar pattern, with increasing risks associated with longer times spent in the ED relative to the 0 to <2 hours reference group. Patients who waited 2 to <4 hours had an aHR of 1.06 (95% CI: 1.04 to 1.07), increasing to an aHR of 1.95 (95% CI: 1.91 to 1.99) after 12 hours spent in the ED.

For models including the ‘immediate’ acuity group, see Supplementary Figure 4. The aHRs for these models show largely the same results as the main models, and do not affect the trend for either the employment or the hospitalisation outcome.

### Secondary analyses

Inclusion of a time-varying hospitalisation indicator in the employment modelling did not change the overall trend, though the aHRs were somewhat attenuated compared with the main analysis (see Figure 4). For example, the aHR at 6 to <8 hours fell from 1.10 to 1.08 (a 20% reduction in excess risk relative to 0 to <2 hours), while that for patients waiting 12+ hours fell from 1.20 to 1.15 (a 25% reduction in excess risk).

**Figure 4.**
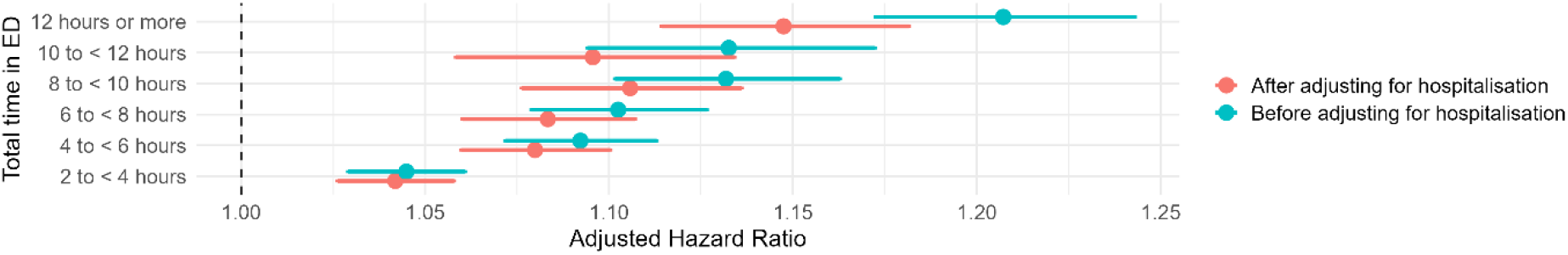
Adjusted hazard ratios for employment before and after adjusting for post-discharge hospitalisation.

## Discussion

### Principal findings

We found that among working-age employees, longer time spent in the ED for non-immediate care was associated with an increased risk of both subsequent loss of employment and future hospitalisation, up to two years after discharge from the ED attendance. For both outcomes, risks rose progressively with increasing ED LOS, demonstrating a graded association.

Individuals experiencing the longest waits, particularly those remaining in the ED for 12 hours or more, had substantially higher hazards of leaving employment and of being hospitalised during follow-up compared with those discharged within two hours of arrival. These patterns were evident in unadjusted cumulative incidence curves and persisted in hazard ratios after adjustment for a wide range of socio-demographic, clinical, and acuity-related factors.

Notably, the risk of leaving employment remained even after adjusting for time-varying hospitalisation, suggesting that longer ED stays may be associated with broader disruptions to health that extend beyond impacts severe enough to warrant subsequent inpatient care. Taken together, these findings indicate that long ED stays may have long-lasting consequences for individuals’ labour market participation and subsequent use of hospital services.

### Research in context

Extended LOS in emergency care has been widely shown to adversely affect clinical outcomes, including increased risks of morbidity and mortality (12,28,32–36). In England, evidence demonstrates a clear relationship between time spent in the ED and mortality risk, with patients experiencing prolonged stays beyond 12 hours facing approximately double the risk of mortality compared with those discharged within two hours (28). Prolonged ED LOS has also been associated with subsequent healthcare utilisation, including an increased likelihood of hospitalisation (36,37). For example, patients remaining in the ED for more than 12 hours have been found to have a significantly greater risk of extended inpatient stays following admission (37). However, these single-centre studies may not be generalisable to the broader emergency patient population, and they only considered hospital inpatient admissions directly following the ED attendance.

Taken together, these findings suggest a potential indirect pathway linking LOS in the ED to labour market outcomes via health deterioration resulting in (extended periods of) hospitalisation. Building on evidence that longer ED stays are associated with an increased likelihood of inpatient admission, separate research has shown that hospitalisation is linked to sustained reductions in earnings and employment status, with per-person losses as large as £18,785 over five years for individuals hospitalised with a stroke (19).

In elective care settings, there is substantial evidence that delays in treatment can influence labour market participation, including effects on earnings, employment status, and work capacity, although findings vary across conditions and contexts (17,18,38,39). In contrast, the literature on emergency care has largely focused on the immediate clinical consequences of prolonged ED stays, with a scarcity of evidence on labour market outcomes.

### Strengths and limitations

To our knowledge, this is the first study to address the evidence gap on the relationship between time spent in the ED and risk of leaving employment. We used individual-level administrative data with near whole-population coverage, integrated across multiple sources (ONS, NHS, HMRC) using linkage at individual level. The maximum follow-up of two years means that our findings are able to shed light on the effect of extended ED stays, rather than just the immediate clinical impacts.

Our study also has limitations. Although we adjusted for a broad range of socio-demographic and health-related factors, it is possible that not all sources of confounding were fully accounted for. We were unable to include measures of ED crowding, hospital staffing or clinical factors that may underpin longer ED stays, such as complex assessments or specialist treatment. Information on patients’ physiological status at presentation, including vital signs, was not available for this study, limiting our ability to fully adjust for clinical severity. As such, the potential for residual confounding remains a key limitation of our analysis, and our findings pertain to statistical associations, not necessarily causal relationships.

Variation in how patient information is recorded across emergency care settings may also affect our findings. In particular, classifications of non-immediate acuity rely on clinician judgement and may not be applied consistently in practice. While acuity can act as an indicator of condition severity, the absence of a standardised approach to its assessment means that coding may differ between sites and clinicians. Given this potential for measurement variation, acuity level was not included as an adjustment in the primary analysis, aside from excluding the “immediate” category and incorporating mode of arrival as a proxy for severity.

The PAYE dataset covers employees only, not the self-employed. Therefore, our study population comprises employees rather than everyone in employment, so our results may not be generalisable to individuals who attend the ED while self-employed. Official labour market data indicate that the self-employed comprise approximately 15% of total employment in the UK (40). In addition, people who stopped being employees during follow-up because they transitioned into self-employment will be misclassified as leaving employment according to our outcome definition. However, such transitions are relatively rare in the UK labour market, typically occurring for less than 0.7% of employees per calendar quarter (40).

Lastly, the generalisability of our findings may be limited by several exclusion criteria applied in the study design. We did not include patients with ED stays exceeding 48 hours, those with missing acuity information, or repeat attendances from individuals with multiple visits during the study period, instead restricting the analysis to first attendances only.

### Conclusion

In conclusion, extended time spent in the ED for non-immediate care is associated with reduced labour market participation and increased healthcare use following discharge among working-age employees in England, which was observed for at least two years after the ED attendance. These findings highlight the possible effects of pressure within emergency care systems that extend beyond the immediate clinical impact, with potential implications for long-term population health, labour supply and economic growth. Further research is needed to explore heterogeneity by occupation type or employment sector, and the impact of extended ED stays on economic outcomes other than employment transitions, such as earnings and benefit receipts.

## Data Availability

The source data used in this study is subject to controlled access due to its sensitive nature and legal constraints.

## Authors’ contributions

HA, DA and VN conceptualised and designed the study. HA, DA prepared the study data and performed the statistical analysis. All authors contributed to interpretation of the results. HA and DA were responsible for the first draft of the manuscript. All authors contributed to critical revision of the manuscript. All authors approved the final manuscript.

## Ethics statement

Ethical approval was obtained from the National Statistician’s Data Ethics Advisory Committee (NSDEC23(18)).

