## Supplementary Materials for "Emergency Department Length of Stay, Employment Status, and Subsequent Risk of Hospitalisation: A survival analysis of linked administrative records in England"

**Supplementary Table 1: Covariates included in models**

| **Variable** | **Coding** | **Source** |
| --- | --- | --- |
| **Age** | Single year of age | ECDS |
| **Sex** | Male, female | Census 2021 |
| **Ethnicity** | White, Asian, Black, Mixed, Other | Census 2021 |
| **Index Multiple Deprivation (IMD) quintile group** | Quintiles of deprivation:  1 (most deprived), 2, 3, 4, 5 (least deprived) | ECDS |
| **Highest qualification** | No qualifications, 1-4 GCSEs/O-levels, 5+ GCSEs/O-levels, apprenticeship, 2+ A-levels or equivalent, degree or above, other qualification, No code required | Census 2021 |
| **ED provider** | Code of hospital attended | ECDS |
| **Chief complaint at arrival** | General / minor / admin, Airway / breathing, Circulation / chest, Environmental, Eye, Gastrointestinal, Genitourinary, Head and neck, Neurological, Obstetrics and gynaecology, Psychosocial / Behaviour change, Skin, Trauma / musculoskeletal | ECDS |
| **Acuity at arrival** | Immediate, Very Urgent, Urgent, Standard, Low | ECDS |
| **Comorbidity: Covid** | Ever, Never | HES APC / OP |
| **Comorbidity: Cancer** | Ever, Never | HES APC / OP |
| **Comorbidity: Diabetes** | Ever, Never | HES APC / OP |
| **Comorbidity: Dementia** | Ever, Never | HES APC / OP |
| **Comorbidity: Serious mental illness** | Ever, Never | HES APC / OP |
| **Comorbidity: Autism** | Ever, Never | HES APC / OP |
| **Comorbidity: Neurological MND, Parkinson’s MS** | Ever, Never | HES APC / OP |
| **Comorbidity: Epilepsy** | Ever, Never | HES APC / OP |
| **Comorbidity: Hypertension** | Ever, Never | HES APC / OP |
| **Comorbidity: Myocardial Infarction** | Ever, Never | HES APC / OP |
| **Comorbidity: Ischaemic hear disease** | Ever, Never | HES APC / OP |
| **Comorbidity: Heart failure** | Ever, Never | HES APC / OP |
| **Comorbidity: Stroke** | Ever, Never | HES APC / OP |
| **Comorbidity: Other respiratory infection** | Ever, Never | HES APC / OP |
| **Comorbidity: Influenza pneumonia** | Ever, Never | HES APC / OP |
| **Comorbidity: Chronic obstructive pulmonary disease and respiratory failure** | Ever, Never | HES APC / OP |
| **Comorbidity: Inflammatory bowel disease** | Ever, Never | HES APC / OP |
| **Comorbidity: Liver disease** | Ever, Never | HES APC / OP |
| **Comorbidity: Rheumatoid arthritis** | Ever, Never | HES APC / OP |
| **Comorbidity: Osteoarthritis** | Ever, Never | HES APC / OP |
| **Comorbidity: Osteoporosis** | Ever, Never | HES APC / OP |
| **Comorbidity: Kidney disease** | Ever, Never | HES APC / OP |

ECDS: Emergency Care Dataset; HES APC / OP: Hospital Episode Statistics Admitted Patient Care / Outpatient.

**Supplementary Table 2. Distributions of covariates by time spent in the ED (<4 hours versus ≥4 hours)**

| **Characteristic** | **Level** | **All (%)** | **< 4 hours (%)** | **≥4 hours (%)** | **Absolute standardised difference (%)** |
| --- | --- | --- | --- | --- | --- |
| **Sex** | Male | 54.7 | 54.2 | 55.4 | 2.4 |
|  | Female | 45.3 | 45.8 | 44.6 |  |
| **Ethnicity** | White | 9.8 | 9.6 | 10.3 | 7.0 |
|  | Asian | 5.1 | 4.9 | 5.5 |  |
|  | Black | 2.2 | 2.2 | 2.2 |  |
|  | Mixed | 2.2 | 2.2 | 2.4 |  |
|  | Other | 80.6 | 81.1 | 79.7 |  |
| **Index Multiple Deprivation (IMD) quintile group** | 1 (most deprived) | 21.5 | 21.4 | 21.6 | 1.0 |
|  | 2 | 21.6 | 21.6 | 21.6 |  |
|  | 3 | 20.2 | 20.2 | 20.2 |  |
|  | 4 | 18.9 | 19.0 | 18.8 |  |
|  | 5 (least deprived) | 17.3 | 17.2 | 17.4 |  |
| **Highest qualification** | Apprenticeship | 4.1 | 4.1 | 4.0 | 4.9 |
|  | Level 1 | 9.6 | 9.5 | 9.9 |  |
|  | Level 2 | 13.8 | 13.7 | 14.0 |  |
|  | Level 3 | 19.5 | 19.6 | 19.3 |  |
|  | Level 4 | 41.8 | 42.3 | 41.1 |  |
|  | No qualifications | 9.2 | 8.9 | 9.6 |  |
|  | Other qualifications | 2.1 | 2.0 | 2.2 |  |
| **Chief complaint at arrival** | Airway / breathing | 4.6 | 3.5 | 6.3 | 57.1 |
|  | Circulation / chest | 15.1 | 13.0 | 18.6 |  |
|  | Environmental | 0.6 | 0.6 | 0.6 |  |
|  | Eye | 2.6 | 3.3 | 1.3 |  |
|  | Gastrointestinal | 11.3 | 8.2 | 16.6 |  |
|  | General / minor / admin | 4.3 | 3.7 | 5.1 |  |
|  | Genitourinary | 2.6 | 2.3 | 3.2 |  |
|  | Head and neck | 3.9 | 4.4 | 3.2 |  |
|  | Neurological | 7.1 | 5.8 | 9.5 |  |
|  | ObGyn | 2.7 | 2.9 | 2.3 |  |
|  | Psychosocial / Behaviour change | 1.4 | 1.0 | 2.0 |  |
|  | Skin | 6.1 | 7.5 | 3.8 |  |
|  | Trauma / musculoskeletal | 29.0 | 34.0 | 20.2 |  |
| **Acuity at arrival** | 2 - Very urgent EC | 7.8 | 5.9 | 11.2 | 104.4 |
|  | 3 - Urgent EC | 40.2 | 34.3 | 50.2 |  |
|  | 4 - Standard EC | 47.2 | 54.3 | 34.9 |  |
|  | 5 - Low acuity EC | 4.8 | 5.5 | 3.7 |  |
| **Comorbidity: COVID-19** | | 0.4 | 0.4 | 0.5 | 2.0 |
| **Comorbidity: Cancer** | | 3.8 | 3.5 | 4.2 | 3.6 |
| **Comorbidity: Dementia** | | 0.0 | 0.0 | 0.0 | 0.2 |
| **Comorbidity: Diabetes** | | 0.4 | 0.3 | 0.5 | 2.9 |
| **Comorbidity: Epilepsy** | | 0.2 | 0.2 | 0.2 | 1.5 |
| **Comorbidity: Heart failure** | | 0.1 | 0.0 | 0.1 | 1.9 |
| **Comorbidity: Hypertension** | | 0.2 | 0.2 | 0.3 | 1.4 |
| **Comorbidity: Inflammatory Bowel Disease** | | 1.3 | 1.2 | 1.6 | 3.4 |
| **Comorbidity: Influenza or pneumonia** | | 0.7 | 0.6 | 0.8 | 2.5 |
| **Comorbidity: Ischaemic hear disease** | | 0.5 | 0.4 | 0.6 | 3.0 |
| **Comorbidity: Kidney disease** | | 1.6 | 1.4 | 1.9 | 4.1 |
| **Comorbidity: Learning disability - Autism** | | 0.0 | 0.0 | 0.0 | 0.0 |
| **Comorbidity: Liver disease** | | 0.2 | 0.2 | 0.3 | 2.4 |
| **Comorbidity: Myocardial infarction** | | 0.4 | 0.4 | 0.5 | 2.8 |
| **Comorbidity: Neurological MND, Parkinson’s Disease and Multiple Sclerosis (MS)** | | 0.1 | 0.1 | 0.1 | 1.1 |
| **Comorbidity: Osteoarthritis** | | 0.9 | 0.9 | 1.0 | 1.3 |
| **Comorbidity: Osteoporosis** | | 0.0 | 0.0 | 0.0 | 0.6 |
| **Comorbidity: Other respiratory infection** | | 1.2 | 1.2 | 1.4 | 1.8 |
| **Comorbidity: Respiratory cancer** | | 0.0 | 0.0 | 0.0 | 1.1 |
| **Comorbidity: Rheumatoid arthritis** | | 0.2 | 0.2 | 0.2 | 1.2 |
| **Comorbidity: Serious mental illness** | | 0.3 | 0.2 | 0.3 | 1.8 |
| **Comorbidity: Stroke** | | 0.2 | 0.2 | 0.3 | 1.9 |

**Supplementary Figure 1: Study sample flow diagram**

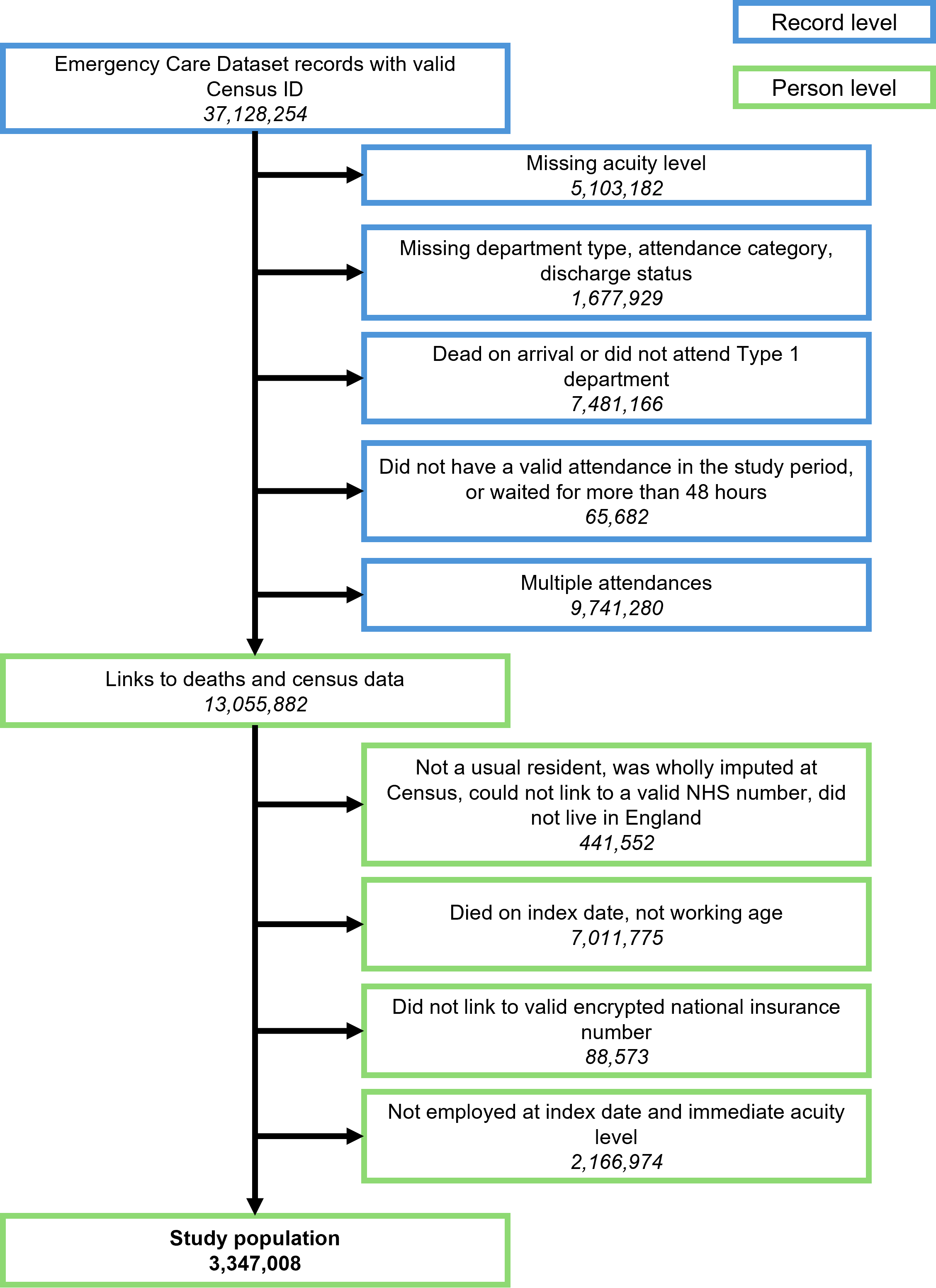

**Supplementary Figure 2: Age- and sex-weighted cumulative incidence curves for leaving employment after discharge from the index ED attendance**

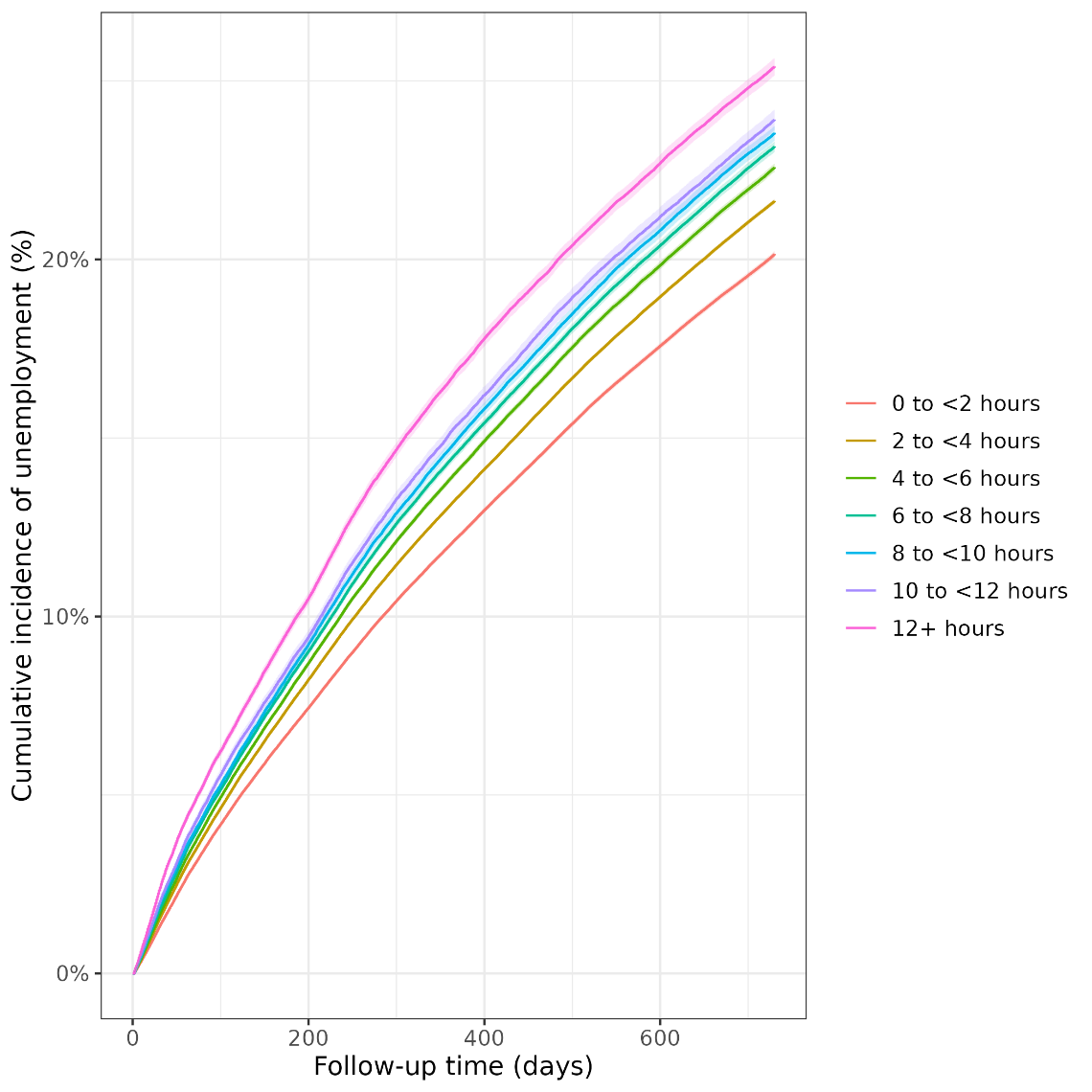

**Supplementary Figure 3: Age- and sex-weighted cumulative incidence curves for being hospitalised after discharge from the index ED attendance**

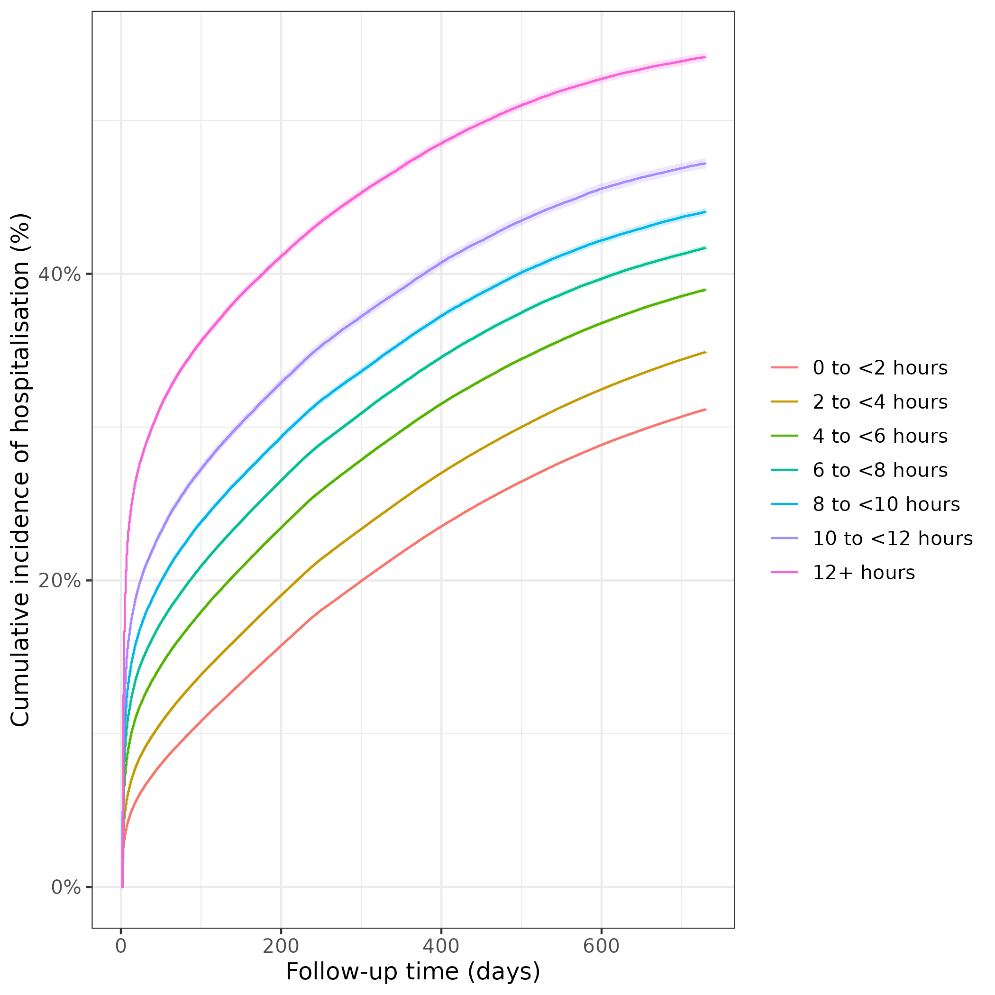

**Supplementary Figure 4: Adjusted hazard ratios for employment from model with ‘immediate’ acuity group included**

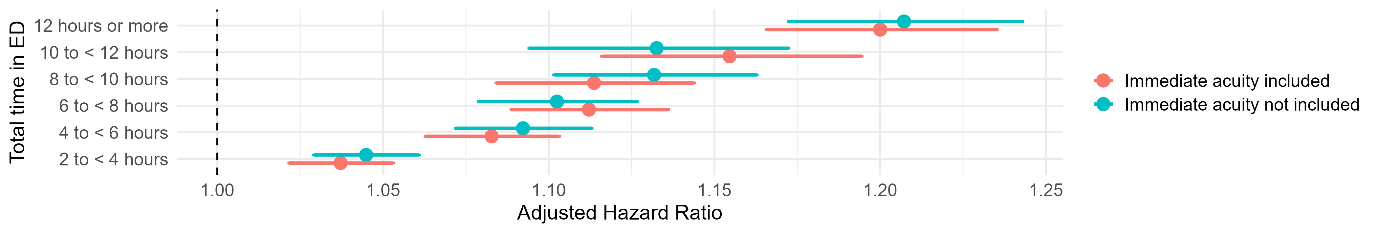

**Supplementary Figure 5: Adjusted hazard ratios for hospitalisation from model with ‘immediate’ acuity group included**

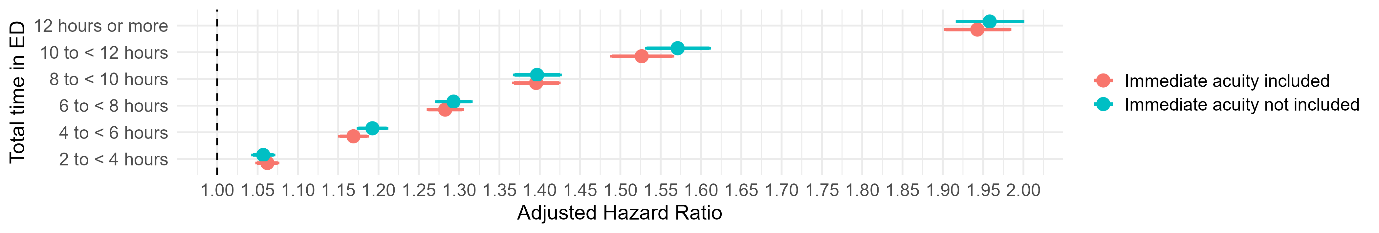
